# Antibiotic coverage in biliary-stented pancreatoduodenectomy: Real-world evidence supporting piperacillin–tazobactam over ampicillin–sulbactam

**DOI:** 10.64898/2026.02.12.26346173

**Authors:** Johannes D Lettner, Palina Matskevich, Carola Focke, Sophia Chikhladze, Stefan Fichtner-Feigl, Stefan Utzolino, Dietrich A Ruess

## Abstract

**Background:** Preoperative biliary stenting alters biliary colonization and may reduce the effectiveness of perioperative antibiotic prophylaxis in pancreatoduodenectomy. Although broader-spectrum regimens have been associated with improved infectious outcomes, their microbiological adequacy in routine clinical practice remains poorly defined. We therefore evaluated the real-world adequacy of a prolonged ampicillin–sulbactam protocol, its association with infectious outcomes and survival, and the potential impact of a universal piperacillin–tazobactam strategy.

**Methods:** We analyzed all consecutive patients who underwent elective pancreatoduodenectomy from 2002 to 2023 at our tertiary center. Demographic, operative, microbiological, and outcome data were retrieved from a prospectively maintained database. Patients were stratified by stent status. Adequacy of prophylaxis was defined as the full in vitro susceptibility of all bile isolates. The outcomes included 30-day infectious morbidity, clinically relevant POPF, PPH, DGE, reoperation, 30- and 90-day mortality and long-term survival. A coverage simulation was performed to compare ampicillin–sulbactam with a hypothetical universal piperacillin–tazobactam. Statistical methods included chi-square/Fisher’s exact tests, Mann–Whitney U tests, Cox models, McNemar’s test and Poisson regression.

**Results:** Of 956 patients, 424 (44%) had a biliary stent. Technical complications were comparable between groups, and rates of POPF and PPH were not increased. However, infectious morbidity was higher in stented patients, including sepsis (RR 1.62, 95% CI 1.05–2.51) and postoperative cholangitis (RR 2.20, 95% CI 1.36–3.56). Thirty- and 90-day mortality were increased (RR 2.88 and 2.73) but lost significance after adjustment. Bile cultures predominantly yielded Enterococcus and Enterobacterales with low ampicillin–sulbactam susceptibility. Overall adequacy was 21.7%. Among patients with bile cultures (n = 474), ampicillin–sulbactam covered 43.7% (207/474) versus 81.2% (385/474) with piperacillin–tazobactam; in stented patients with cultures (n = 397), coverage increased from 41.8% to 78.1%. Adequate ampicillin–sulbactam coverage was not associated with reduced infectious outcomes in Poisson models.

**Conclusion:** Preoperative stenting creates a polymicrobial, partially resistant biliary niche that ampicillin–sulbactam does not sufficiently cover. Our data shows that a piperacillin–tazobactam strategy substantially improves coverage and was therefore implemented at our center.

**Core message:**

- Stented patients exhibit a distinct infectious risk profile characterized by Enterococcus-and Enterobacterales-dominated bile colonization rather than increased rates of technical complications.
- In stented patients, real-world microbiological coverage of ampicillin–sulbactam was limited, and in vitro susceptibility did not independently translate into reduced postoperative infectious morbidity.
- Broader prophylaxis, such as piperacillin/tazobactam, aligns with the actual flora and nearly doubles theoretical coverage, addressing the mismatch between stent-associated biofilms and narrow regimens.

## Introduction

Infectious complications are a significant cause of early morbidity following pancreatoduodenectomy, and they are the focus of ongoing research (1).

Patients with preoperative biliary drainage are particularly vulnerable among all risk groups. Biliary stents become colonized rapidly and almost invariably, leading to polymicrobial contamination. This increases the risk of postoperative wound infections, intra-abdominal abscesses, cholangitis, sepsis, and other serious complications (2–4).

Accumulating evidence shows that the microbiology of patients with stents differs fundamentally from that of patients without stents. In this context, alternative drainage strategies such as surgical biliary drainage have been proposed to potentially reduce ascending microbial contamination of the biliary tree compared with endoscopic stenting or percutaneous transhepatic drainage, although these approaches are not widely adopted (5).

Metagenomic analyses of biliary stents reveal consistent colonization by Enterococcus faecalis and Enterobacterales, which includes Escherichia coli, Klebsiella pneumoniae, and Enterobacter cloacae. This colonization is often accompanied by mixed flora of aerobes and anaerobes, as well as non-fermenters, including Pseudomonas aeruginosa and Stenotrophomonas maltophilia (6,7). These organisms exhibit heterogeneous susceptibility patterns and harbor antimicrobial resistance genes, necessitating adequate antibiotic treatment during surgery (8–10). This raises concerns about the adequacy of traditional perioperative prophylaxis in this setting. Against this backdrop, the adequacy of narrower-spectrum prophylactic regimens in stented patients is increasingly being questioned, particularly in light of evolving stent-associated biliary flora. Recent randomized trials investigating perioperative prophylaxis in pancreatic surgery suggest that broader antimicrobial coverage is necessary (3,11–13). However, systematic evaluations of prophylaxis adequacy against patient-specific bile isolates are scarce (14). Additionally, the clinical consequences of inadequate coverage in the context of preoperative biliary stenting remain unclear (3,15). This study had three aims: to evaluate the appropriateness of Ampicillin/Sulbactam prophylaxis based on susceptibility in pancreatoduodenectomy, to simulate the expected improvement in coverage under a universal piperacillin-tazobactam strategy, and to systematically examine the clinical consequences of inadequate coverage, including postoperative morbidity, recurrence of the same pathogen, and survival.

## Methods

### Study design

This was a retrospective, single-center cohort study conducted at a high-volume pancreatic surgery center. All consecutive patients who underwent pancreatoduodenectomy between 2002 and 2023 were screened. According to the institutional protocol, perioperative prophylaxis consisted of intravenous ampicillin–sulbactam (Unacid). In patients with preoperative biliary stenting, prophylaxis was routinely prolonged to 72 hours. Piperacillin–tazobactam (Pip/Taz) was occasionally administered in presumed high-risk cases at the discretion of the treating surgeon. The study protocol complied with the Declaration of Helsinki and was approved by the local ethics committee (reference 25-1294-S1-retro). Informed consent was waived due to the retrospective design and use of anonymized data. Reporting followed the STROBE (Strengthening the Reporting of Observational Studies in Epidemiology) guidelines.

### Participants

Eligible patients were adults (18 years or older) undergoing elective pancreatoduodenectomy with documented perioperative prophylaxis and available pre- or intraoperative bile cultures. Those with missing microbiological data, palliative procedures, or incomplete records were excluded. For all primary analyses, the cohort was stratified by biliary stent status. Since patients with preoperative biliary stents have a distinct microbiological exposure profile, all appropriateness- and coverage-based analyses were restricted to patients with endoscopic ERC stents. Patients with PTCD or surgical biliary drainage were not included in this subgroup analysis. Non-stented patients served as the biological reference group.

### Surgical procedures

Pancreatic head resections were performed as classical Whipple procedures (distal gastrectomy), pylorus-preserving pancreatoduodenectomy (PPPD), or pylorus-resecting pancreatoduodenectomy (PRPD) .Standardized reconstruction followed, consisting of duct-to-mucosa pancreaticojejunostomy or pancreatogastrostomy, hepaticojejunostomy, and duodenojejunostomy or gastrojejunostomy. One or two drains were routinely placed adjacent to the pancreatic anastomosis. Minimally invasive pancreatoduodenectomy was performed as either a hybrid laparoscopic or a fully robotic procedure. Minimally invasive surgery was chosen based on preoperative imaging, comorbidity, and patient history. It was generally avoided in cases of borderline resectable disease or after neoadjuvant therapy. No additional selection criteria were applied because conversion to an open procedure was possible at any time. Conversion was defined as an open approach initiated before dissecting the pancreas along with the superior mesenteric artery. The hybrid minimally invasive technique has been described previously (16). In brief, resection was performed with five trocars placed in the upper abdomen. After the pancreatic head was resected, an approximately 8-cm midline or subcostal incision was made to remove the specimen. Reconstruction was then performed through this incision.

### Intraoperative stent management, bile sampling, and antibiotic regimen

At our center, all stents were completely removed during pancreatoduodenectomy in patients with preoperative biliary drainage. Immediately after dividing the bile duct, a sterile bile sample was obtained and submitted for aerobic and anaerobic culture, including full susceptibility testing. Perioperative prophylaxis consisted of intravenous ampicillin–sulbactam administered before incision. In patients with preoperative biliary stenting, prophylaxis was routinely prolonged for 72 hours. Postoperative antibiotic adjustments were permitted once susceptibility results became available. At the time of protocol implementation, the previous German S3 guideline on peri-interventional and perioperative antibiotic prophylaxis recommended cefazolin plus metronidazole for most abdominal procedures. The use of ampicillin–sulbactam therefore represented an expanded-spectrum strategy, particularly in combination with prolonged prophylaxis in stented patients. However, increasing resistance rates among Enterobacterales, particularly *Escherichia coli*, have since led to a re-evaluation of ampicillin–sulbactam in hepatopancreatobiliary surgery. The updated German S3 guideline (effective December 2024) now recommends piperacillin–tazobactam for pancreatoduodenectomy based on accumulating evidence and resistance data(17). For coverage simulation, all isolates were re-evaluated under a hypothetical universal piperacillin–tazobactam regimen.

### Data collection

Data were obtained from prospectively maintained institutional databases and validated through a systematic review of charts. The extracted variables included demographic and clinical characteristics, operative details, stent-related parameters, microbiological findings from preoperative bile cultures, perioperative antibiotic prophylaxis, and postoperative outcomes, including infectious and noninfectious morbidity, reoperation, and 30- and 90-day mortality and overall, one- and five-year survival.

### Inclusion/Exclusion criteria

Eligible patients were adults who underwent elective pancreatoduodenectomy between 2002 and 2023 and had complete clinical documentation, including detailed records of preoperative and perioperative antibiotic prophylaxis. For analyses involving preoperative biliary stenting, only patients with documented stent placement and available preoperative bile cultures were included, as well as those with full microbiological and susceptibility data. Patients were excluded if they lacked complete perioperative antibiotic documentation; had no bile cultures despite stent placement; had missing microbiological or susceptibility data; or underwent palliative procedures or non-standard resections.

### Clinical Outcome definitions

The adequacy of the administered perioperative prophylaxis, as defined by complete in vitro susceptibility of all isolated bile pathogens to the given regimen, was assessed based on coverage. The primary clinical endpoint was 30-day infectious morbidity, which was defined as a composite of surgical-site infection, intra-abdominal infection, pneumonia, or sepsis. Each component was adjudicated according to predefined clinical and microbiological criteria (Table 2). The secondary endpoints included the recurrence of identical pathogens within 30 days, clinically relevant postoperative pancreatic fistulas (grades B and C according to the International Study Group of Pancreatic Surgery (ISGPS)), post-pancreatectomy hemorrhage (ISGPS), reoperation within 30 days, and all-cause of mortality at 30 and 90 days. Overall survival (OS) was defined as the interval from the date of surgery to death from any cause, and survivors were censored at the last available follow-up.

### Coverage-based evaluation and coverage simulation

The adequacy of the administered perioperative prophylaxis was defined as the complete in vitro susceptibility of all pathogens isolated from an individual’s intraoperative bile culture

Microbiological susceptibility testing yielded categorical results classified as susceptible (S), intermediate (I), or resistant (R) according to standard laboratory criteria. To coverage analysis, only isolates categorized as susceptible were considered covered; intermediate and resistant results were classified as non-coverage. In addition to the coverage profile of the administered prophylactic regimen, the theoretical activity of alternative antibiotics, including piperacillin–tazobactam, was evaluated. Antibiotic classes were grouped into first-, second- and last-line categories based on microbiological susceptibility patterns. To quantify the potential benefit of broader perioperative prophylaxis strategies, a counterfactual coverage simulation was performed. This model contrasted the observed coverage achieved with ampicillin–sulbactam with the hypothetical coverage expected if all patients had received piperacillin–tazobactam and incorporated maximal achievable coverage using a stepwise escalation framework. Differential coverage (Δ%) was defined as the proportion of patients fully covered under the universal piperacillin–tazobactam scenario minus the proportion fully covered under the administered ampicillin–sulbactam regimen.

### Statistical analysis

The study had a retrospective, full-census design, so no priori sample size calculation was performed. Continuous variables are presented as median (interquartile range [IQR]) and were compared using the Mann–Whitney U test. Categorical variables were analyzed using Pearson’s chi-squared test or Fisher’s exact test, depending on the expected cell counts. Baseline characteristics: Adjusted effect estimates for postoperative complications were derived from multivariable logistic regression models. Survival outcomes were analyzed using Kaplan-Meier estimators with log-rank testing and Cox proportional hazards models. One-year and five-year survival probabilities were obtained from fixed time points on the time-to-event scale. The proportional hazards assumptions and model diagnostics were routinely assessed.

Multivariable regression models were used to adjust for relevant baseline differences. Relative risks were estimated using Poisson regression with robust standard errors (Table 9). Separate models were fitted for the ampicillin–sulbactam–treated stent subgroup and the full stent cohort, in both unadjusted and adjusted forms. Poisson regression was prespecified for outcomes with non-rare incidence, as odds ratios may overestimate the risk in these cases. Antibiotic coverage adequacy was defined by bile culture susceptibility. We assessed real-world appropriateness and within-patient paired comparisons between ampicillin–sulbactam and hypothetical universal piperacillin–tazobactam prophylaxis using McNemar’s test with exact two-sided p values. Microbiological findings are summarized descriptively, and the full isolated panel is provided in Supplementary Table S1. All analyses were conducted using R version 4.5.2 and IBM SPSS Statistics version 31.0. Statistical significance was defined as P < 0.05 (two-sided).

### Covariates in multivariable models

Multivariable models were adjusted for the following variables: age, sex, gender, American Society of Anesthesiologists grade (ASA), neoadjuvant therapy, final histopathology (pancreatic ductal adenocarcinoma (PDAC) versus non-PDAC), and preoperative bilirubin concentration. Due to significant skewness, bilirubin was entered as a transformed continuous variable.

### Recurrence of the same pathogen

The recurrence of the same pathogen was evaluated by matching preoperative bile isolates with postoperative microbiological cultures. Same-pathogen recurrence was defined as the re-isolation of at least one organism with an identical species and susceptibility profile from a normally sterile site. This occurred predominantly in blood cultures or, when available, in intra-abdominal specimens within 30 days after surgery.

## Results

### Study cohort and characteristics

A total of 1.019 pancreatoduodenectomies were performed at our center from 2002 to 2023. Of those, 956 patients met all the eligibility criteria and were included in the final analysis. Patients were excluded primarily due to missing microbiological or perioperative documentation. Specifically, 25 patients with stents lacked intraoperative bile cultures, 12 patients had incomplete susceptibility data, 22 patients had insufficiently documented perioperative antibiotic prophylaxis, and four patients had incomplete clinical records. Among the patients included, 424 (44.4%) had a preoperative biliary stent, while 532 (55.6%) did not (Table 1). The median age differed significantly between the two groups (68.5 vs. 65.0 years, p = 0.002). The sex distribution (41.9% vs. 47.9% female, p = 0.068), BMI, and diabetes rates were similar. A higher proportion of non-stent patients had an American Society of Anesthesiologists (ASA) class of III or higher (62.4% vs. 46.7%, p < 0.001). Most comorbidities, including pulmonary, cardiac, hepatic, and alcohol-related conditions, were comparable. Renal comorbidity was more prevalent in the non-stent group (11.8% vs. 7.8%, p = 0.040). There was a trend toward higher rates of nicotine abuse in non-stented patients (p = 0.054). Neoadjuvant therapy and serum amylase levels were similar between the two groups. Preoperative icterus (81.8% vs. 23.3%, p < 0.001) and serum bilirubin levels (median 1.65 vs. 0.50 mg/dL, p < 0.001) were markedly higher in stented patients, reflecting the need for biliary drainage. Among stented patients, the median dwell time was 24.5 days, and 53.3% had a dwell time longer than 30 days. The surgical approach was similar between the two groups, with comparable rates of open and minimally invasive procedures (p = 0.618). Final histopathological diagnoses differed significantly (p = 0.040). Pancreatic ductal adenocarcinoma (PDAC) was more prevalent in stented patients (78.5% vs. 45.3%), while intraductal papillary mucinous neoplasm (IPMN), neuroendocrine tumor (NET), chronic pancreatitis, and other pathologies were more frequent in non-stented patients (Table 1).

**Table 1.** Characteristics of total collective. Abbreviations: BMI, body mass index; ASA, American Society of Anesthesiologists physical status classification; PPPD, pylorus-preserving pancreatoduodenectomy; PDAC, pancreatic ductal adenocarcinoma; IPMN, intraductal papillary mucinous neoplasm; NET, neuroendocrine tumour.

|  | All n = 956 | Stent = 424 | No Stent = 532 | p |
| --- | --- | --- | --- | --- |
| **Demographic factors** |  |  |  |  |
| Age, years, median [IQR] | 67.0 [58.0–74.0] | 68.5 [60.3–74.3] | 65.0 [55.0–73.0] | **0.002** |
| Sex, female, n [%] | 433 [45.3] | 178 [41.9] | 255 [47.9] | 0.068 |
| BMI, kg/m<sup>2</sup>, median [IQR] | 24.7 [22.3–27.3] | 24.3 [22.0–26.7] | 24.6 [22.2–26.9] | 0.109 |
| ASA ≥ III, n [%] | 430 [45.0] | 198 [46.7] | 332 [62.4] | **<0.001** |
| Diabetes mellitus, yes, n [%] | 136 [14.2] | 65 [15.3] | 71 [13.4] | 0.402 |
| Exocrine insufficiency, yes, n [%] | 43 [4.5] | 15 [3.5] | 28 [5.3] | 0.213 |
| **Comorbidities, yes, n [%]** |  |  |  |  |
| Lungs, yes, n [%] | 149 [15.6] | 75 [17.7] | 74 [13.9] | 0.127 |
| Heart, yes, n [%] | 97 [10.1] | 48 [11.3] | 49 [9.2] | 0.284 |
| Kidneys, yes, n [%] | 96 [10.0] | 33 [7.8] | 63 [11.8] | 0.040 |
| Liver, yes, n [%] | 179 [18.7] | 84 [19.8] | 95 [17.9] | 0.454 |
| Nicotin abuse, yes, n [%] | 249 [26.1] | 97 [22.9] | 152 [28.6] | 0.054 |
| Alcohol abuse, yes, n [%] | 132 [13.8] | 62 [14.6] | 70 [13.2] | 0.571 |
| Neoadjuvant therapy, yes, n [%] | 24 [2.5] | 13 [3.1] | 11 [2.1] | 0.406 |
| Serum Amylase, U/L, median [IQR] | 28 [17–48] | 28 [14.5–52.3] | 28.5 [19–42.3] | 0.372 |
| **Biliary stenting** |  |  |  |  |
| Duration of stenting, days, median [IQR] | 29.0 [20.0–49.0] | 24.5 [17–32.3] | - |  |
| Stent dwelling time ≤ 30 days, yes, n [%] | 199 [20.8] | 199 [46.9] | - |  |
| Stent dwelling time >30 days, yes, n [%] | 226 [23.6] | 226 [53.3] | - |  |
| Preoperative icterus, yes, n [%] | 471 [49.3] | 347 [81.8] | 124 [23.3] | **<0.001** |
| Serum bilirubin, mg/dl, median [IQR] | 0.7 [0.4–2.0] | 1.65 [0.88–3.73] | 0.50 [0.40–1.20] | **<0.001** |
| **Surgical procedure** |  |  |  |  |
| Open PPPD, n [%] | 675 [70.6] | 303 [71.5] | 372 [69.9] | **0.618** |
| Minimal invasive, n [%] | 281 [29.4] | 121 [28.5] | 160 [30.1] | — |
| **Postoperative** |  |  |  |  |
| Histopathology |  |  |  | **0.040** |
| PDAC, n [%] | 574 [60.1] | 333 [78.5] | 241 [45.3] |  |
| IPMN, n [%] | 99 [10.4] | 4 [0.9] | 95 [17.8] |  |
| NET, n [%] | 49 [5.1] | 8 [1.9] | 41 [7.7] |  |
| Chronic pancreatitis, n [%] | 110 [11.5] | 36 [8.5] | 74 [13.9] |  |
| Others, n [%] | 124 [13.0] | 43 [10.1] | 83 [15.6] |  |

### Postoperative outcomes, complications, and mortality

Antibiotic regimens differed substantially between groups (see Table 2). Patients with biliary stents received ampicillin-sulbactam significantly more often (48.3%) than non-stented patients (1.9%, p < 0.001). Cephalosporin-based prophylaxis was less common in the stent group (25.5% vs. 69.5%, p < 0.001). There were also group differences in fluoroquinolone-based regimens and “other last-line” regimens (p < 0.001 and p = 0.048, respectively). Piperacillin–tazobactam and carbapenem-based prophylaxis were used infrequently and did not differ significantly between groups. When administered, broader-spectrum regimens were selected on the basis of preoperative microbiological data and known resistance patterns. Overall, postoperative complications occurred within 30 days in 62.4% of the total cohort, with comparable rates in the stent group (60.8%) and the non-stent group (63.7%, p = 0.810). POPF occurred in 40.6% of patients overall and was less prevalent in patients with a stent (35.6%) than in those without (44.5%, p = 0.003). Clinically relevant POPF (CR-POPF) occurred in 18.7% of patients overall (16.0% in the stent group vs. 20.9% in the non-stent group, p = 0.011). Postpancreatectomy hemorrhage (PPH) occurred in 11.4% of patients (9.0% in the stent group vs. 13.3% in the non-stent group, p = 0.030). The rate of bile leak (8.1% overall) did not differ significantly between groups (p = 0.066). Delayed gastric emptying occurred in 10.6% of patients overall (10.4% in the stent group and 10.7% in the non-stent group, p = 0.400). Severe complications (Clavien-Dindo ≥ III) were documented in 35.2% of patients, with lower rates in the stent group: (31.1%) than in the non-stent group (38.5%, p = 0.028). Surgical complications were comparable across groups. Anastomotic insufficiency occurred in 31.0% of patients overall. There were lower rates in the stent group (27.4%) than in patients without a stent (33.8%, p = 0.032). Abdominal wound dehiscence was uncommon and did not differ between groups (2.4% total; p = 0.540; see Table 2). Postoperative organ dysfunction was uncommon. Acute kidney injury occurred in 4.0% of patients overall, with no difference between those with stents (3.3%) and those without (4.5%, p = 0.360). Sepsis developed in 7.6% of patients overall, but it was significantly more frequent in the stent cohort (9.7%) than in the non-stent cohort (6%, p = 0.046) (Table 2). Several significant differences were observed in infectious complications (Table 2). Abdominal wound infections occurred in 15.3% of patients overall, with higher rates in stented patients (17.9%) than in patients without stents (13.0%, p = 0.049). Intra-abdominal abscesses were less common in stented patients (11.1%) than in non-stented patients (18.2%, p = 0.003). Postoperative cholangitis predominantly occurred in the stent group (5.0% vs. 0.9%, p < 0.001). Pneumonia occurred in 5.5% of patients overall and was less prevalent in the stent group (3.8% vs. 7.0%, p = 0.022). Urinary tract infections occurred in 5.9% of patients overall and were less common in the stent group (4.0% vs. 7.3%, p = 0.041). Mortality differed between the cohorts: Thirty- and ninety-day mortality were significantly higher in the stented group (both p < 0.001) (Table 2). Reoperations within 30 days were required for 15.4% of the total cohort. Lower rates were seen in the stent group (11.8%) than in the non-stent group (18.2%, p = 0.006). Length of hospital and ICU stays did not differ between groups. Overall, 30-day mortality was 7.0%, substantially higher in the stent group (12.7% vs. 2.4%, p < 0.001). The pattern was identical for 90-day mortality (17.5% vs. 6.4%, p < 0.001). One-year survival rates were similar between the two groups (around 81%-82%, p = 0.505). Five-year overall survival was 42.6% for the entire cohort, which was significantly lower for stented patients (27.1%) than for those without stents (56.1%, p < 0.001) (Figure 1).

**Figure 1.**
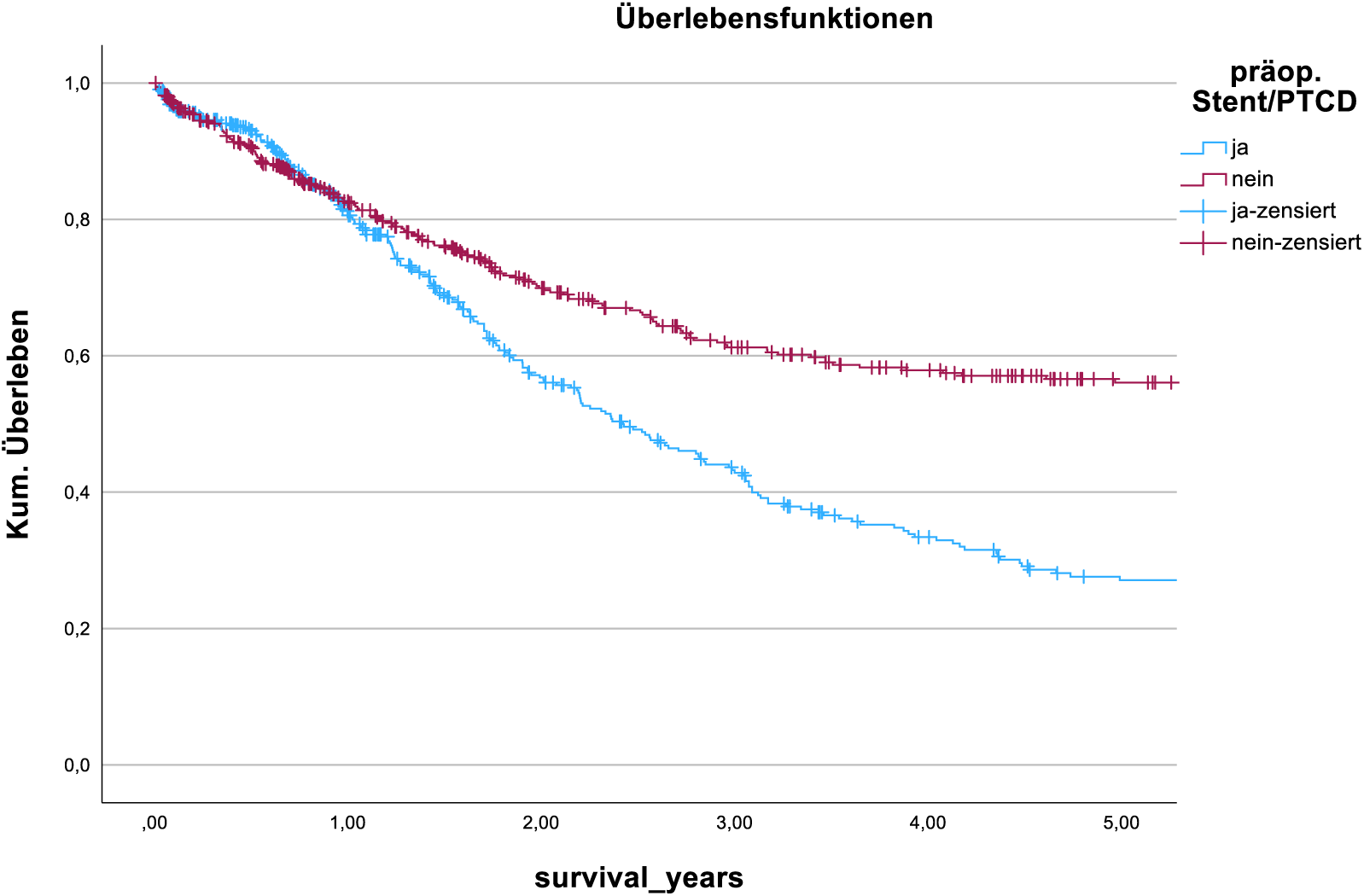
Kaplan–Meier curves of overall survival stratified by preoperative biliary stenting. Blue line, patients with a preoperative biliary stent; red line, patients without stent. Censoring is indicated by crosses. At 1 year, overall survival was similar in both groups (80.6% in stented vs 82.4% in non-stented patients), but the curves separate progressively thereafter, with a clearly lower survival probability in the stent group by 2 years and beyond. Log-rank p < 0.001.

**Table 2.** Antibiotic prophylaxis, Morbidity, complications and mortality. Abbreviations: POPF, postoperative pancreatic fistula; CR-POPF, clinically relevant postoperative pancreatic fistula; PPH, postpancreatectomy hemorrhage; DGE, delayed gastric emptying; AKI, acute kidney injury; KDIGO, Kidney Disease: Improving Global Outcomes; ICU, intensive care unit.

|  | All n = 956 | Stent = 424 | Non Stent = 532 | p |
| --- | --- | --- | --- | --- |
| **Antibiotic prophylaxis regimens** |  |  |  |  |
| Ampicillin/Sulbactam, yes, n [%] | 215 [22.5] | 205 [48.3] | 10 [1.9] | **<0.001** |
| Cephalosporin-based, yes, n [%] | 565 [59.1] | 108 [25.5] | 457 [85.9] | **<0.001** |
| Piperacillin/Tazobactam, yes, n [n%] | 18 [1.9 %] | 9 [2.1] | 9 [1.7] | 0.470 |
| Fluoroquinolone-based, yes, n [n%] | 122 [12.7] | 83 [19.6] | 39 [7.3] | **<0.001** |
| Carbapenem-based, yes, n [%] | 7 [0.7] | 4 [0.9] | 3 [0.6] | 0.070 |
| Other last line regimens, yes, n [%] | 26 [2.7] | 17 [4.0] | 9 [1.7] | **0.048** |
| **Overall Morbidity** |  |  |  |  |
| Any Complications within 30d, yes, n [%] | 597 [62.4] | 258 [60.8] | 339 [63.7] | 0.810 |
| POPF, n [%] | 388 [40.6] | 151 [35.6] | 237 [44.5] | **0.003** |
| CR-POPF | 179 [18.7] | 68 [16.0] | 111 [20.9] | **0.011** |
| PPH, n [%] | 109 [11.4] | 38 [9.0] | 71 [13.3] | **0.030** |
| B & C, n [%] | 77 [8.1] | 28 [6.6] | 49 [9.2] | 0.066 |
| DGE, n [%] | 101 [10.6] | 44 [10.4] | 57 [10.7] | 0.400 |
| Clavien-Dindo ≥ III, n [%] | 337 [35.2] | 132 [31.1] | 205 [38.5] | **0.028** |
| **Surgical complications** |  |  |  |  |
| Anastomosis insufficiency, any yes, n [%] | 296 [31] | 116 [27.4] | 180 [33.8] | **0.032** |
| Abdominal wound dehiscence, yes, n [%] | 23 [2.4] | 12 [2.8] | 11 [2.1] | 0.540 |
| **Postoperative Organ dysfunction** |  |  |  |  |
| AKI defined according to KDIGO, yes, n [%] | 38 [4.0] | 14 [3.3] | 24 [4.5] | 0.360 |
| Sepsis, yes, n [%] | 73 [7.6] | 41 [9.7] | 32 [6] | **0.046** |
| **Infectious complications** |  |  |  |  |
| Abdominal wound infection, yes, n [%] | 146 [15.3] | 76 [17.9] | 69 [13.0] | **0.049** |
| Intra-abdominal abscess, yes, n [%] | 144 [15.1] | 47 [11.1] | 97 [18.2] | **0.003** |
| Postoperative cholangitis, yes, n [%] | 26 [2.7] | 21 [5.0] | 5 [0.9] | **<0.001** |
| Postoperative pneumonia, yes, n [%] | 53 [5.5] | 16 [3.8] | 37 [7.0] | **0.022** |
| Urinary tract infection, yes, n [%] | 56 [5.9] | 17 [4.0] | 39 [7.3] | **0.041** |
| **Mortality** |  |  |  |  |
| Revision <30d, yes, n [%] | 147 [15.4] | 50 [11.8] | 97 [18.2] | **0.006** |
| Hospital stays, days, median [IQR] | 15 [12-22] | 14.5 [12-20.5] | 15 [11-22.3] | 0.623 |
| ICU stay, days, median [IQR] | 4 [3-9] | 7 [3-9] | 4 [4-6] | 0.297 |
| 30-day mortality | 67 [7.0] | 54 [12.7] | 13 [2.4] | **<0.001** |
| 90-day mortality | 108 [11.3] | 74 [17.5] | 34 [6.4] | **<0.001** |
| 1-year overall survival, % [CI] | 81.6 [78.9-84.3] | 80.6 [76.2-84.4] | 82.4 [78.9-85.9] | 0.505 |
| 5-year overall survival, % [CI] | 42.6 [38.5-46.7] | 27.1 [21.6-32.6] | 56.1 [50.8-61.4] | **<0.001** |

### Crude Risk Ratios

The crude risk ratios comparing stented patients (n = 424) and non-stented patients (n = 532) are summarized in Table 3. The incidence of any complication within 30 days was similar between the two groups (60.8% vs. 63.7%; RR: 0.95, CI: 0.86–1.06). Rates of postoperative pancreatic fistula (POPF) (35.6% vs. 44.5%; RR: 0.80, CI: 0.68–0.94) and clinically relevant POPF (16.0% vs. 20.9%; RR: 0.77, CI: 0.58–1.01) were lower in the stented group. The incidence of PPH (9.0% vs. 13.3%; RR: 0.68, CI: 0.44–1.02), bile leak (6.6% vs. 9.2%; RR: 0.72, CI: 0.46–1.12), and delayed gastric emptying (10.4% vs. 10.7%; RR: 0.97, CI: 0.67–1.41) did not differ significantly. Clavien-Dindo grade ≥ III occurred less frequently in the stent group (31.1% vs. 38.5%; RR: 0.81, CI: 0.68–0.97). Anastomotic insufficiency was less common in stented patients (27.4% vs. 33.8%; RR: 0.81, CI: 0.67–0.98). There was no relevant difference in abdominal wound dehiscence (2.8% vs. 2.1%; RR: 1.37, CI: 0.61–3.07). The rates of acute kidney injury were comparable (3.3% vs. 4.5%; RR: 0.73, CI: 0.38–1.40). However, sepsis occurred more frequently in stented patients (9.7% vs. 6.0%; RR: 1.62, CI: 1.05–2.51). Stented patients had higher rates of abdominal wound infection (17.9% vs. 13%; RR: 1.38, CI: 1.02–1.87). Intra-abdominal abscesses occurred at similar frequencies (11.1% vs. 9.2%; RR: 1.20, CI: 0.80–1.82). Postoperative cholangitis occurred more frequently in the stent group (5.0% vs. 0.9%; RR: 2.20, CI: 1.36–3.56). There were differences in rates of postoperative pneumonia (3.8% vs. 1.9%; RR: 1.95, CI: 0.83–4.57) and urinary tract infection (4.0% vs. 7.3%; RR: 0.55, CI: 0.31–0.95). Revision within 30 days was less common in stented patients (11.8% vs. 18.2%; RR: 0.65, CI: 0.47–0.89). The stent group had a higher thirty-day mortality rate (12.7% vs. 2.4%; RR 5.21, CI 2.88–9.42). Similarly, 90-day mortality increased (17.5% vs. 6.4%; RR: 2.73, CI: 1.86–4.02).

**Table 3.**
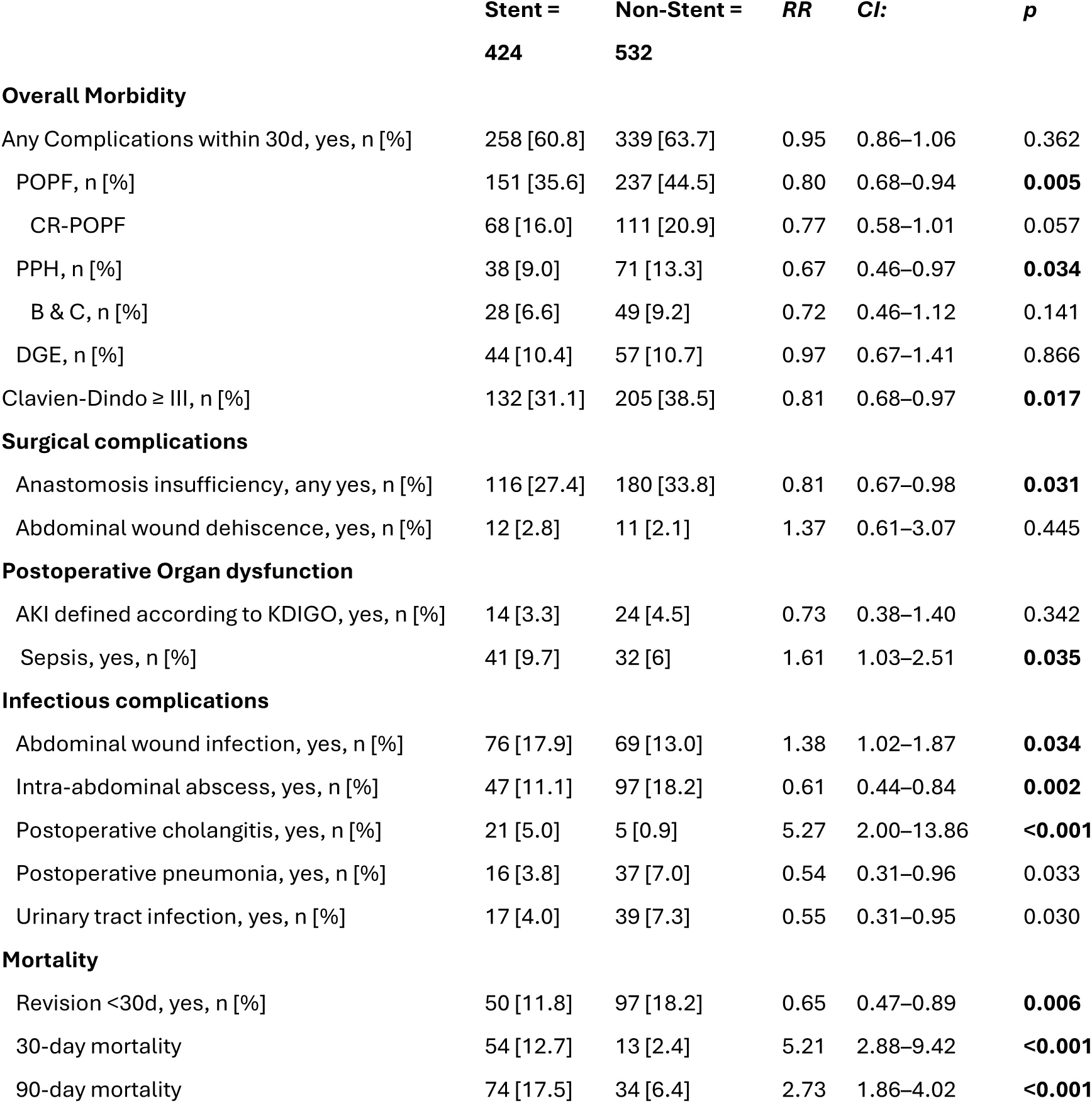
Crude Risk Ratios (RR) for Stent versus Non-Stent Cohort. Abbreviations: POPF, postoperative pancreatic fistula; CR-POPF, clinically relevant postoperative pancreatic fistula; PPH, postpancreatectomy hemorrhage; DGE, delayed gastric emptying; AKI, acute kidney injury; KDIGO, Kidney Disease: Improving Global Outcomes; ICU, intensive care unit.

|  | **Stent =**<br>**424** | **Non-Stent =**<br>**532** | **RR** | **CI:** | **p** |
| --- | --- | --- | --- | --- | --- |
| **Overall Morbidity** |  |  |  |  |  |
| Any Complications within 30d, yes, n [%] | 258 [60.8] | 339 [63.7] | 0.95 | 0.86–1.06 | 0.362 |
| POPF, n [%] | 151 [35.6] | 237 [44.5] | 0.80 | 0.68–0.94 | **0.005** |
| CR-POPF | 68 [16.0] | 111 [20.9] | 0.77 | 0.58–1.01 | 0.057 |
| PPH, n [%] | 38 [9.0] | 71 [13.3] | 0.67 | 0.46–0.97 | **0.034** |
| B & C, n [%] | 28 [6.6] | 49 [9.2] | 0.72 | 0.46–1.12 | 0.141 |
| DGE, n [%] | 44 [10.4] | 57 [10.7] | 0.97 | 0.67–1.41 | 0.866 |
| Clavien-Dindo ≥ III, n [%] | 132 [31.1] | 205 [38.5] | 0.81 | 0.68–0.97 | **0.017** |
| **Surgical complications** |  |  |  |  |  |
| Anastomosis insufficiency, any yes, n [%] | 116 [27.4] | 180 [33.8] | 0.81 | 0.67–0.98 | **0.031** |
| Abdominal wound dehiscence, yes, n [%] | 12 [2.8] | 11 [2.1] | 1.37 | 0.61–3.07 | 0.445 |
| **Postoperative Organ dysfunction** |  |  |  |  |  |
| AKI defined according to KDIGO, yes, n [%] | 14 [3.3] | 24 [4.5] | 0.73 | 0.38–1.40 | 0.342 |
| Sepsis, yes, n [%] | 41 [9.7] | 32 [6] | 1.61 | 1.03–2.51 | **0.035** |
| **Infectious complications** |  |  |  |  |  |
| Abdominal wound infection, yes, n [%] | 76 [17.9] | 69 [13.0] | 1.38 | 1.02–1.87 | **0.034** |
| Intra-abdominal abscess, yes, n [%] | 47 [11.1] | 97 [18.2] | 0.61 | 0.44–0.84 | **0.002** |
| Postoperative cholangitis, yes, n [%] | 21 [5.0] | 5 [0.9] | 5.27 | 2.00–13.86 | **<0.001** |
| Postoperative pneumonia, yes, n [%] | 16 [3.8] | 37 [7.0] | 0.54 | 0.31–0.96 | 0.033 |
| Urinary tract infection, yes, n [%] | 17 [4.0] | 39 [7.3] | 0.55 | 0.31–0.95 | 0.030 |
| **Mortality** |  |  |  |  |  |
| Revision <30d, yes, n [%] | 50 [11.8] | 97 [18.2] | 0.65 | 0.47–0.89 | **0.006** |
| 30-day mortality | 54 [12.7] | 13 [2.4] | 5.21 | 2.88–9.42 | **<0.001** |
| 90-day mortality | 74 [17.5] | 34 [6.4] | 2.73 | 1.86–4.02 | **<0.001** |

### Adjusted Logistic Regression

The rate of POPF was lower in stented patients (aOR: 0.660, CI: 0.580–0.751, p < 0.001).The risk of PPH was also reduced (aOR: 0.594, CI: 0.389–0.907, p = 0.016). There was a non-significant association with Clavien–Dindo grade ≥ III (aOR 0.782, CI: 0.421–0.982, p = 0.052). The adjusted odds of sepsis did not differ significantly (aOR: 1.563; CI: 0.956–2.554; p: 0.075). There was no significant difference in abdominal wound infection (aOR 1.374, CI 0.958–1.972, p = 0.085). Intra-abdominal abscesses occurred less frequently in the stent group (p < 0.001). Cholangitis occurred more frequently in stented patients (aOR: 4.811; CI: 1.787–12.956; p = 0.002). Revision within 30 days occurred less frequently (aOR: 0.567; CI: 0.390–0.825; p = 0.003). Thirty-day and ninety-day mortality were significantly higher in the stent group (Table 4).

**Table 4.** Adjusted Logistic Regression. Abbreviations: POPF, postoperative pancreatic fistula; PPH, postpancreatectomy hemorrhage.

| Postoperative Outcome | aOR | CI: | *p* |
| --- | --- | --- | --- |
| POPF | 0.660 | 0.580-0.751 | **<0.001** |
| PPH | 0.594 | 0.389-0.907 | **0.016** |
| Clavien-Dindo ≥ III | 0.782 | 0.421-0.982 | 0.052 |
| Anastomosis insufficiency |  |  |  |
| Sepsis | 1.563 | 0.956-2.554 | 0.075 |
| Abdominal wound infection | 1.374 | 0.958-1.972 | 0.085 |
| Intra-abdominal abscess | 0.516 | 0.351-0.758 | **<0.001** |
| Cholangitis | 4.811 | 1.787-12.956 | **0.002** |
| Revision <30d | 0.567 | 0.390-0.825 | **0.003** |
| 30-day mortality | 3.569 | 1.939-6.568 | **<0.001** |
| 90-day mortality | 1.979 | 1.288-3.040 | **0.002** |

### Survival Analysis

In the unadjusted analysis, thirty-day mortality was higher (HR: 3.397, CI: 1.824–6.327, p < 0.001). However, after adjustment, the association reversed (HR: 0.627, CI: 0.514–0.765, p < 0.001). Ninety-day mortality exhibited a comparable trend, demonstrating an elevated unadjusted hazard ratio (HR 1.859, CI: 1.237–2.794, p = 0.003) and a decreased adjusted hazard ratio (HR 0.589, CI: 0.508–0.755, p < 0.001). Unadjusted overall survival was lower in stented patients (HR: 0.544; CI: 0.455–0.674; p < 0.001), but the adjusted model showed no significant difference (HR: 1.087; CI: 0.911–1.297; p = 0.354) (Table 5).

**Table 5.** Survival Analysis for stented vs non-stented patients.

| Postoperative Outcome | Model | HR | 95% CI | *p* |
| --- | --- | --- | --- | --- |
| 30-day mortality | Unadjusted Cox | 3.397 | 1.824-6.327 | **<0.001** |
|  | Adjusted Cox | 0.627 | 0.514-0.765 | **<0.001** |
| 90-day mortality | Unadjusted Cox | 1.859 | 1.237-2.794 | **0.003** |
|  | Adjusted Cox | 0.589 | 0.508-0.755 | **<0.001** |
| Overall Survival | Unadjusted Cox | 0.544 | 0.455-0.674 | **<0.001** |
|  | Adjusted Cox | 1.087 | 0.911-1.297 | 0.354 |

### Microbiological spectrum

Among patients with biliary stents, the ten most frequently isolated organisms from preoperative bile cultures were Enterococcus faecalis (n = 224), Enterococcus faecium (n = 115), Escherichia coli (n = 104), and Enterobacter cloacae (n = 89). The next most frequent isolates were Klebsiella oxytoca (n = 66), Klebsiella pneumoniae (n = 65), Streptococcus anginosus group (n = 65), Citrobacter freundii (n = 45), Proteus vulgaris (n = 27), and Hafnia alvei (n = 23) (Table 6). A detailed list of all isolated organisms, including low-frequency species, anaerobes, and Gram-positive rods, is provided in Supplementary Table S1.

**Table 6.**
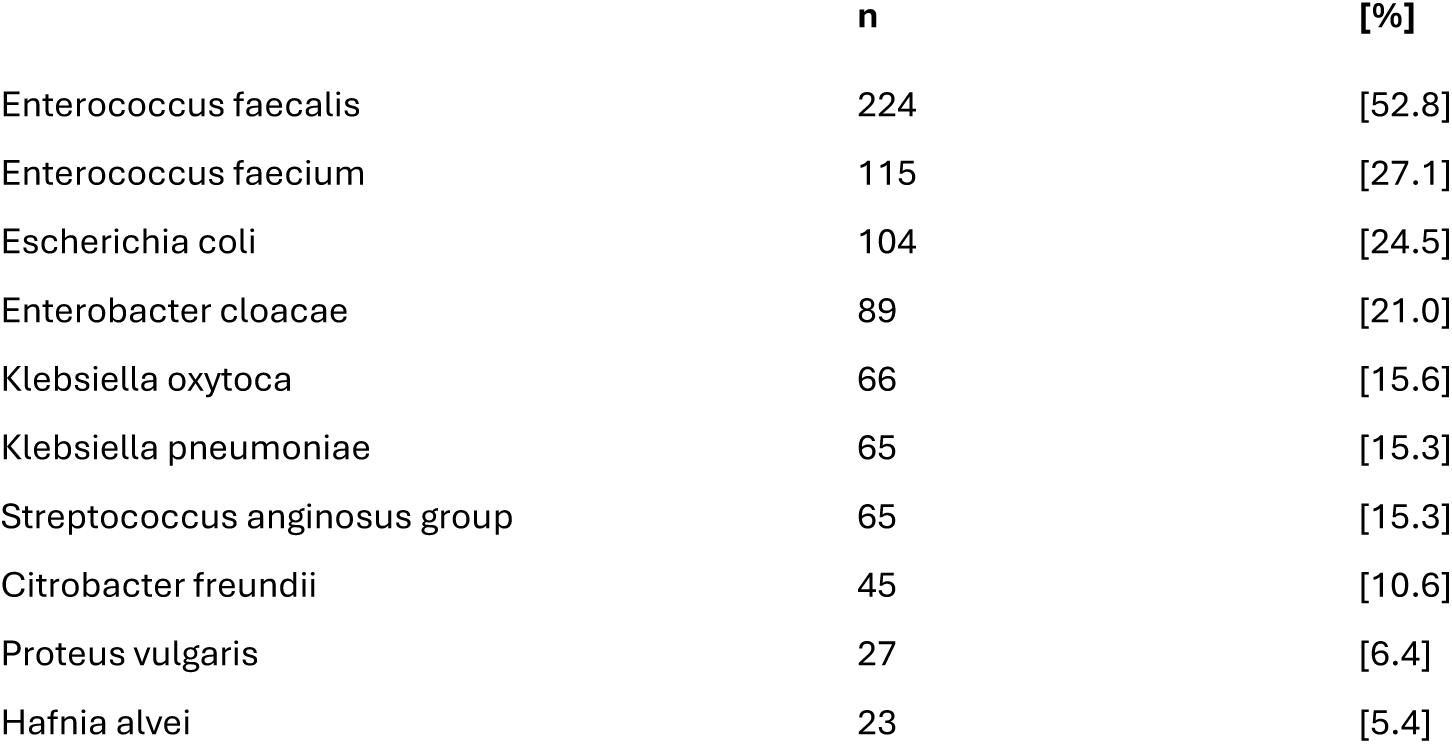
Top 10 isolated organisms from preoperative bile cultures in patients with biliary stents. Multiple organisms per patient were possible. The full microbiological spectrum, including low-frequency isolates and anaerobes, is provided in Supplementary Table S1.

|  | n | [%] |
| --- | --- | --- |
| *Enterococcus faecalis* | 224 | [52.8] |
| *Enterococcus faecium* | 115 | [27.1] |
| *Escherichia coli* | 104 | [24.5] |
| *Enterobacter cloacae* | 89 | [21.0] |
| *Klebsiella oxytoca* | 66 | [15.6] |
| *Klebsiella pneumoniae* | 65 | [15.3] |
| *Streptococcus anginosus* group | 65 | [15.3] |
| *Citrobacter freundii* | 45 | [10.6] |
| *Proteus vulgaris* | 27 | [6.4] |
| *Hafnia alvei* | 23 | [5.4] |

### Susceptibility-based appropriateness of perioperative prophylaxis

As shown in Table 7, susceptibility-based appropriateness differed markedly between groups. Ampicillin-sulbactam would have provided complete in vitro coverage for 207 out of 956 patients (21.7%), with notably higher coverage among stent recipients (39.1% versus 7.7%; p < 0.001). Piperacillin/tazobactam would have been adequate only for isolates not covered by ampicillin-sulbactam and occurred more frequently in stented patients (36.0% vs. 12.2%; p < 0.001). These values reflect the regimen-specific requirement profile. The separate coverage simulation in Table X quantifies the overall theoretical coverage achievable under a universal piperacillin/tazobactam strategy. Requirements for carbapenem, fluoroquinolone, and last-line antibiotics were also predominantly confined to the stent cohort (all p < 0.001). Same-pathogen recurrence occurred in 129 patients (13.5%), and it was more prevalent among stented patients (25.2% vs. 4.1%; p < 0.001).

**Table 7.** Theoretical coverage of perioperative antibiotic regimens based on in-vitro bile culture susceptibilities and same-pathogen recurrence. Definitions: Same-pathogen recurrence within 30 days refers to postoperative identification of the same bacterial species as in the preoperative bile culture within 30 days after surgery.

|  | All = 956 | Stent = 424 | Non-Stent = 532 | *p* |
| --- | --- | --- | --- | --- |
| With ampicillin/sulbactam, n [%] | 207 [21.7] | 166 [39.1] | 41 [7.7] | **<0.001** |
| Piperacillin/tazobactam required, n [%] | 218 [22.8] | 153 [36] | 65 [12.2] | **<0.001** |
| Carbapenem required, n [%] | 35 [3.7] | 34 [8] | 1 [0.2] | **<0.001** |
| Fluoroquinolone required, n [%] | 17 [1.8] | 16 [3.8] | 1 [0.2] | **<0.001** |
| Last-line antibiotics required, n [%] | 44 [4.6] | 41 [9.6] | 3 [0.6] | **<0.001** |
| Same pathogen recurrence within 30 days, n [%] | 129 [13.5] | 107 [25.2] | 22 [4.1] | **<0.001** |

### Coverage Simulation

Of the 474 patients with available preoperative or intraoperative bile cultures, real-world prophylaxis with Ampicillin/Sulbactam provided appropriate microbial coverage in 207 patients (43.7%). In a hypothetical scenario using piperacillin/tazobactam, coverage increased to 385 patients (81.2%), corresponding to an absolute improvement of 37.5% (Table 8). Within the stent subgroup (n = 397), appropriateness increased from 166 patients (38.3%) under the real-world regimen to 320 patients (80.6%) under the hypothetical regimen—an improvement of 38.8%. In the non-stent subgroup (n = 77), appropriateness increased from 41 patients (53.3%) to 65 patients (84.4%), corresponding to a 31.2% improvement. All comparisons showed statistically significant differences (p < 0.001) (Table 8).

**Table 8.** Coverage simulation of patients with pre- or intraoperative bile culture.

|  | **Real-world appropriateness with**<br>**ampicillin/sulbactam** | **Appropriateness with**<br>**piperacillin/tazobactam** | **Δ (%)** | **p** |
| --- | --- | --- | --- | --- |
| Total (n = 474) | 207 [43.7] | 385 [81.2] | +37.5 | **<0.001** |
| Stent (n = 397) | 166 [38.3] | 320 [80.6] | +38.8 | **<0.001** |
| No Stent (n = 77) | 41 [53.3] | 65 [84.4] | +31.2 | **<0.001** |

### Poisson Regression

The Poisson regression results for the ampicillin/sulbactam-treated stent cohort (n = 205) and the full stent cohort (n = 424) are summarized in Table 9. Neither unadjusted nor adjusted models showed statistically significant associations for sepsis, cholangitis, abdominal wound infection, revision within 30 days, 30-day mortality, 90-day mortality, or overall survival (all p > 0.05). Abdominal wound infection was the only outcome with a significant association in the full cohort, observed in both the unadjusted (RR: 1.09, CI: 1.03–1.16, p = 0.002) and adjusted models (RR: 1.10, CI: 1.04–1.17, p = 0.002). All other endpoints, including sepsis, cholangitis, revision within 30 days, 30-day mortality, 90-day mortality, and overall survival, showed no significant associations in either model (all p > 0.05).

**Table 9.** Poisson regression for ampicillin/sulbactam treated patients only of the stent cohort and of full stent cohort.

| **Ampicillin-sulbactam treated**<br>**stent patients ( n =205)** | **Model** | **RR** | **CI:** | **p** |
| --- | --- | --- | --- | --- |
| Sepsis | Unadjusted Poisson | 1.00 | 0.93-1.08 | 0.979 |
|  | Adjusted Poisson | 0.99 | 0.93-1.06 | 0.922 |
| Cholangitis | Unadjusted Poisson | 0.97 | 0.89-1.09 | 0.725 |
|  | Adjusted Poisson | 0.98 | 0.88-1.08 | 0.648 |
| Abdominal wound infection | Unadjusted Poisson | 1.10 | 0.99-1.22 | 0.064 |
|  | Adjusted Poisson | 1.10 | 0.99-1.23 | 0.088 |
| Revision <30d | Unadjusted Poisson | 1.00 | 0.90-1.11 | 0.905 |
|  | Adjusted Poisson | 0.99 | 0.89-1.09 | 0.808 |
| 30-day mortality | Unadjusted Poisson | 0.96 | 0.95-0.99 | 0.987 |
|  | Adjusted Poisson | 0.97 | 0.94-0.99 | 0.986 |
| 90-day mortality | Unadjusted Poisson | 0.69 | 0.21-6.10 | 0.737 |
|  | Adjusted Poisson | 0.85 | 0.08-9.64 | 0.899 |
| Overall Survival | Unadjusted Poisson | 1.09 | 0.63-1.88 | 0.768 |
|  | Adjusted Poisson | 0.95 | 0.61-1.50 | 0.839 |
| **Full stent cohort ( n = 424)** |  |  |  |  |
| Sepsis | Unadjusted Poisson | 1 | 0.97-1.03 | 0.953 |
|  | Adjusted Poisson | 1.01 | 0.97-1.04 | 0.770 |
| Cholangitis | Unadjusted Poisson | 1.03 | 0.99-1.06 | 0.127 |
|  | Adjusted Poisson | 1.03 | 0.99-1.07 | 0.122 |
| Abdominal wound infection | Unadjusted Poisson | 1.09 | 1.03-1.16 | **0.002** |
|  | Adjusted Poisson | 1.10 | 1.04-1.17 | **0.002** |
| Revision <30d | Unadjusted Poisson | 1.02 | 0.98-1.05 | 0.365 |
|  | Adjusted Poisson | 1.02 | 0.98-1.06 | 0.387 |
| 30-day mortality | Unadjusted Poisson | 1.79 | 0.41-10.46 | 0.518 |
|  | Adjusted Poisson | 1.48 | 0.24-9.12 | 0.676 |
| 90-day mortality | Unadjusted Poisson | 0.95 | 0.26-3.44 | 0.943 |
|  | Adjusted Poisson | 0.72 | 0.18-2.88 | 0.639 |
| Overall Survival | Unadjusted Poisson | 1.21 | 0.91-1.61 | 0.180 |
|  | Adjusted Poisson | 1.15 | 0.87-1.53 | 0.315 |

## Discussion

The present study shows that preoperative bile duct drainage is associated with a significant shift in the postoperative risk profile after pancreatoduodenectomy. While surgical complications such as POPF, PPH, and anastomotic leakage occurred more frequently in the non-stent group, the stent group showed a clearly increased risk of infection with more sepsis, wound infections, and especially cholangitis, which is reflected in increased 30- and 90-day mortality (18,19).

### Stent-Associated Vulnerability: An infectious, not just technical, phenomenon

Both groups were comparable in terms of demographics and clinical characteristics. Higher bilirubin levels and more frequent jaundice were indications for drainage. Despite theoretical concerns about local inflammation, previous studies found no consistent increase in technical complications due to preoperative stents (4,20). Our analysis also showed no increase in technical risks. Even though contaminated bile was associated with POPF in individual studies, this was not confirmed in our cohort. In fact, the opposite was true (21,22). Therefore, the vulnerability of the stent group is primarily infectious in nature, even after adjusting for factors such as age, jaundice, ASA, neoadjuvant therapy and tumour biology. The increased rates of cholangitis and sepsis are consistent with studies describing the rapid, dense colonisation of stents (23–26). Metagenomic analyses provide a more complete picture by detecting a wide range of resistance genes and factors associated with biofilms (3,6). Taken together, these factors create a robust microbiological niche that is difficult to address therapeutically, thereby reinforcing the limitations of narrow prophylaxis regimens.

### Postoperative complications, risk and survival analyses

There was no increase in surgical complications in the stent group; POPF (ISGPS), PPH and anastomotic leaks were even less frequent. In contrast, infectious morbidity was clearly increased, particularly sepsis, postoperative cholangitis, and wound infections. Intra-abdominal abscesses and reoperations were more prevalent in the non-stent group, consistent with its higher POPF rates. However, early mortality was concentrated in the stent group. The adjusted models confirm this pattern: Biliary drainage was not an independent driver of surgical complications, and lower POPF, PPH and revision rates were observed even after controlling relevant covariates. The only stable effect was the persistently increased risk of postoperative cholangitis. After adjustment, sepsis lost its significance, reflecting the higher baseline burden in the stent group. However, the unadjusted signal remains clinically relevant.

Survival analyses demonstrate significant confounding of the raw data. The increased early mortality disappeared completely after adjustment and was even slightly reversed — indicating that the raw signal primarily reflects jaundice burden, ASA status, and oncological selection. Earlier studies (Sahora, van der Gaag, Scheufele et al.) also found no causal relationship between stenting and mortality, but documented a clear relationship between stenting, biliary colonization, and infectious complications (18,21,27,28). Our data is consistent: the increased unadjusted risk is not explained by the stent itself, but by polymicrobiology, resistance shifts and biofilm-associated tolerance. The observed difference in 5-year overall survival must be interpreted cautiously. This late divergence reflects oncological selection rather than any effect of stenting or infectious complications. As long-term survival after pancreatoduodenectomy is primarily determined by tumour biology, the 5-year signal should not be over-attributed to perioperative factors.

### Microbiological context and implications for prophylaxis

The microbiological profile of the stent group closely matched the patterns reported in large RCTs: dominance of Enterococcus species and a broad range of Enterobacterales, consistent with Ellis et al. In our cohort, E. faecalis and E. faecium were the leading isolates, accompanied by diverse Gram-negative rods. This reproducible spectrum illustrates the limitations of narrow β-lactam regimens and explains the increased infectious morbidity observed in stented patients. A key contribution of this work is the direct link between this flora and the appropriateness of prophylaxis. Ampicillin-sulbactam continues to be used routinely in Europe but is only partially effective against stent-associated organisms. The actual appropriateness was correspondingly low; this is consistent with recent studies documenting increasing polymicrobiology (3). Poisson regression analysis revealed that ‘adequate’ in-vitro coverage did not necessarily translate into clinical protection, as neither sepsis nor cholangitis, nor early mortality or revisions, were reduced. The lack of benefit may suggest that in-vitro susceptibility does not reflect true in-vivo activity in stent-associated polymicrobial biofilms. Considering the lack of clinical response despite in-vitro ‘adequacy’, the significantly higher resistance load in contaminated bile remains crucial. Enterococcus species exhibit intrinsic cephalosporin resistance and diminishing ampicillin sensitivity, while Enterobacterales subvert traditional treatment regimens via AmpC and ESBL mechanisms (3,7,15). In conjunction with biofilm-induced tolerance, a resistance architecture emerges that systematically overwhelms narrow prophylactic measures.

### Simulation of broader prophylaxis and potential clinical implications

The coverage simulation showed that piperacillin/tazobactam addresses the structural limitations of narrow regimens: theoretical coverage nearly doubled and covered the dominant Enterococcus and Enterobacterales isolates. This does not imply a direct clinical outcome benefit but highlights the structural mismatch between stent-associated flora and currently used prophylaxis. Randomized studies support this interpretation: D’Angelica et al. showed a reduction in postoperative SSIs under broader prophylaxis (10). Ellis et al. demonstrated the superiority of piperacillin/tazobactam in cefoxitin-resistant biliary pathogens, precisely the organisms that dominate in our stent cohort (3,15).

### Strengths and limitations

The strengths include the large cohort, the prospectively maintained high-volume database, systematic intraoperative bile sampling and the integration of culture-based microbiology with detailed clinical endpoints. The Poisson models enabled robust, clinically interpretable risk estimation, while the paired simulation design reduced interindividual differences in sensitivity patterns. Limitations include the retrospective, single-centre design; missing cultures in part of the non-stent group; incomplete recording of biofilm-associated tolerance; and the lack of clinical validation of the simulated strategy. A formal MDR/ESBL classification was not consistently available throughout the study period due to changes in laboratory standards and test panels. Therefore, the analysis focused on clinically relevant susceptibility to standard perioperative regimens. POPF rates are multifactorial in origin and can only be interpreted to a limited extent (21,22,29). The long study period also reflects historical changes in perioperative standards, particularly antibiotic regimens.

## Conclusions

Our data show that preoperative biliary stent implantation leads to complex, polymicrobial colonization that narrow perioperative regimens do not adequately cover. The coverage simulation is consistent with the results of randomized controlled trials (RCTs) demonstrating the superiority of piperacillin/tazobactam. Considering the clear shift towards resistant Enterobacterales and Enterococcus species, as well as the heterogeneity of stent-associated flora, it seems appropriate and ethically justifiable to provide this high-risk group with broader perioperative prophylaxis. Based on this data and the available evidence, prophylaxis at our center has consistently switched to piperacillin/tazobactam.

## Data Availability

All data is available from the corresponding authors upon reasonable request.

## Acknowledgements

J.D. Lettner was supported by the Postdoctoral Research Funding Program of the Research Commission of the Faculty of Medicine, University of Freiburg. D.A. Ruess is supported by the German Research Foundation Deutsche Forschungsgemeinschaft; CRC1479 P17; Project ID: 441891347 and by the German Cancer Aid, Deutsche Krebshilfe; Project ID: 70113697. The funders played no role in study design, data collection, analysis and interpretation of data, or the writing of this manuscript. No additional external funding was received for the conduct or publication of this work.

## Authors contributions

J.D.L. contributed to conceptualization, methodology, formal analysis, investigation, data curation, writing – original draft, writing – review and editing, supervision, and project administration. P.M. contributed to investigation, data curation, and writing – review and editing. C.F. contributed to writing – review and editing. S.C. contributed to writing – review and editing and supervision. S.F.-F. contributed to writing – review and editing. S.U. contributed to conceptualization and writing – review and editing. D.A.R. contributed to conceptualization, methodology, writing – review and editing, supervision, and project administration. All authors read and approved the final version of the manuscript.

