## Supplemental Material for "Antibiotic coverage in biliary-stented pancreatoduodenectomy: Real-world evidence supporting piperacillin–tazobactam over ampicillin–sulbactam"

**Table S1. Comprehensive bile culture spectrum in patients with preoperative biliary stenting (n = 424).** The table lists all isolated microorganisms, grouped by Gram-negative Enterobacterales, non-fermenters, Gram-positive cocci, anaerobes/low-virulence flora, and other Gram-positive rods. Multiple isolates per patient were possible.

| | **n** | **[%]** |
|---|---|---|
| **Gram-negative Enterobacterales** | | |
| Escherichia coli | 104 | [24.5] |
| Enterobacter cloacae | 89 | [21.0] |
| Klebsiella oxytoca | 66 | [15.6] |
| Klebsiella pneumoniae | 65 | [15.3] |
| Citrobacter freundii | 45 | [10.6] |
| Hafnia alvei | 23 | [5.4] |
| Proteus vulgaris | 27 | [6.4] |
| Proteus mirabilis | 7 | [1.7] |
| Morganella morganii | 11 | [2.6] |
| Serratia marcescens | 8 | [1.9] |
| Citrobacter koseri | 14 | [3.3] |
| Citrobacter amalonaticus | 2 | [0.5] |
| Citrobacter braakii | 1 | [0.2] |
| Klebsiella aerogenes | 1 | [0.2] |
| Klebsiella variicola | 1 | [0.2] |
| Enterobacter aerogenes | 10 | [2.4] |
| Enterobacter agglomerans | 1 | [0.2] |
| Enterobacter spp. | 3 | [0.7] |
| Serratia fonticola | 2 | [0.5] |
| Serratia liquefaciens | 2 | [0.5] |
| Providencia rettgeri | 1 | [0.2] |
| Haemophilus parainfluenzae | 2 | [0.5] |
| Pasteurella spp. | 1 | [0.2] |
| **Non-fermenters** | | |
| Pseudomonas aeruginosa | 9 | [2.1] |
| Pseudomonas citronellolis | 1 | [0.2] |
| Stenotrophomonas maltophilia | 1 | [0.2] |
| **Gram-positive cocci** | | |
| Enterococcus faecalis | 224 | [52.8] |
| Enterococcus faecium | 115 | [27.1] |
| Streptococcus anginosus group | 65 | [15.3] |
| Enterococcus casseliflavus | 17 | [4.0] |

| | | |
|---|---|---|
| Enterococcus avium | 16 | [3.8] |
| Staphylococcus epidermidis | 13 | [3.1] |
| Staphylococcus aureus | 12 | [2.8] |
| Enterococcus gallinarum | 12 | [2.8] |
| Streptococcus viridans group | 13 | [3.1] |
| Streptococcus sanguinis | 14 | [3.3] |
| Enterococcus durans | 10 | [2.4] |
| Streptococcus gordonii | 7 | [1.7] |
| Staphylococcus coagulase-negative | 7 | [1.7] |
| Streptococcus intermedius | 5 | [1.2] |
| Streptococcus mitis | 3 | [0.7] |
| Streptococcus spp. | 3 | [0.7] |
| Enterococcus hirae | 3 | [0.7] |
| Streptococcus bovis | 2 | [0.5] |
| Staphylococcus haemolyticus | 2 | [0.5] |
| Streptococcus parasanguinis | 2 | [0.5] |
| Streptococcus constellatus | 2 | [0.5] |
| Streptococcus pneumoniae | 1 | [0.2] |
| Streptococcus salivarius | 1 | [0.2] |
| Enterococcus malodoratus | 1 | [0.2] |
| Staphylococcus hominis | 1 | [0.2] |
| Lactococcus garvieae | 2 | [0.5] |
| **Anaerobes / low-virulence flora** | | |
| Clostridium perfringens | 21 | [5.0] |
| Clostridium spp. | 3 | [0.7] |
| Clostridium bifermentans | 2 | [0.5] |
| Clostridium sordellii | 1 | [0.2] |
| Bacteroides spp. | 1 | [0.2] |
| Bacteroides fragilis | 1 | [0.2] |
| Veillonella parvula | 2 | [0.5] |
| Lactobacillus spp. | 4 | [0.9] |
| Lactobacillus plantarum | 2 | [0.5] |
| Lactobacillus paracasei | 1 | [0.2] |
| Lactobacillus reuteri | 1 | [0.2] |
| Bifidobacterium spp. | 1 | [0.2] |
| **Other Gram-positive rods** | | |
| Actinomyces turicensis | 1 | [0.2] |
| Corynebacterium spp. | 1 | [0.2] |
